# Antithrombotic Therapy After Transcatheter Aortic Valve Implantation in Patients with Long-Term Indication for Anticoagulation: Systematic Review and Meta-Analysis

**DOI:** 10.1101/2020.09.22.20198119

**Authors:** Gonçalo Costa, José Pedro Sousa, Lino Gonçalves, Rogério Teixeira

**Affiliations:** Serviço de Cardiologia, Centro Hospitalar e Universitário de Coimbra, Coimbra, Portugal; Faculdade de Medicina da Universidade de Coimbra, Coimbra, Portugal; Coimbra Institute for Clinical and Biomedical Research (iCBR), Coimbra, Portugal

**Keywords:** Atrial Fibrillation, Aortic Stenosis, TAVI, Anticoagulant therapy

## Abstract

**Objective:** The aim of this study was to compare antithrombotic regimens following transcatheter aortic valve implantation (TAVI) in patients requiring long-term oral anticoagulation (OAC).

**Methods:** We systematically searched PubMed, Embase, and Cochrane databases for interventional and observational studies comparing OAC to OAC plus single antiplatelet therapy (SAPT).

**Results:** Five studies were included (four registry-based and one randomized controlled trial), comprising a total of 1318 patients. Our meta-analysis revealed lower rates of severe bleeding (pooled odds ratio [OR] 0.46 [0.31, 0.69], P<0.01, I^2^=0%) and major bleeding (pooled OR 0.46 [0.27,0.79], P<0.01, I^2^=0%) for the OAC group than for the OAC-plus-SAPT group. There was a nonsignificant trend towards reduced life-threatening bleeding events in the OAC group (pooled OR 0.54 [0.27,1.08], P=0.08, I^2^=11%). There was no difference between groups in the risks of stroke (pooled OR 1.02 [0.58,1.80], P=0.58, I^2^= 0%) or all-cause mortality (pooled OR 1.04 [0.75,1.42], P=0.83, I^2^= 0%) after TAVI.

**Conclusions:** Our pooled analysis suggests that for patients with an indication for long-term OAC after TAVI, double anti-thrombotic therapy, compared to OAC alone, increased the risk of bleeding without reducing cerebrovascular events and all-cause mortality.

**Key questions:**

- **What is already known about this subject?** Transcatheter Aortic Valve Implantation (TAVI) is a landmark techinique in interventional cardiology with growing clinical experience and continuous refinement of procedural techniques and devices. Nevertheless, the post-TAVI antithrombotic regimen guidelines are largely based on expert opinions and followed variably across the globe.
- **What does this study add?** Our pooled analysis suggests that for patients with an indication for long-term anticoagulation after TAVI, double anti-thrombotic therapy, compared to OAC alone, increased the risk of bleeding without reducing cerebrovascular events and all-cause mortality.
- **How might this impact on clinical practice?** Our finding may prove insightful for future recommendations regarding the conundrum of the best antithrombotic strategy, particularly for patients with AF.

## INTRODUCTION

Transcatheter aortic valve implantation (TAVI) is used in high-risk patients with symptomatic severe aortic stenosis (AS)(1). The procedure is complicated by major and life-threatening bleeding in 3% to 13% of patients, and the incidental rate of stroke after TAVI varies from 1% to 12%(2,3). Atrial fibrillation (AF) is the most common sustained arrhythmia in the general population. Its prevalence increases with age, affecting more than 10% of people aged ≥ 80 years, and, overall, the prevalence and incidence of AF are growing worldwide(4,5). AF is an indication for long-term oral anticoagulation (OAC) with a vitamin K antagonist (VKA) or a direct-acting oral anticoagulant (DOAC)(4,5). AF and AS share multiple common risk factors, including age and hypertension, and AS itself is associated with a higher rate of AF because of pressure overload imposed on the left atrium(6). Optimal peri-TAVI management of antithrombotic therapy represents a relevant issue for patients with AF. Current practice recommendations on antithrombotic treatment for patients requiring OAC after TAVI suggest the use of VKA alone(7) or in combination with aspirin or clopidogrel(8). The rationale for the additional therapy is to reduce the risk of thromboembolic complications, but, in patients without a long-term indication for anticoagulation, two randomized trials have not shown a benefit of clopidogrel plus aspirin therapy in terms of reducing ischemic and vascular events following TAVI(9,10). Moreover, a trend towards an increased risk of bleeding was noted for patients on dual antiplatelet therapy(10). Additionally, the recent POPular-TAVI trial cohort B showed that OAC alone was associated with reduced bleeding compared to OAC with clopidogrel in post-TAVI patients requiring long-term OAC(11).

The purpose of this study was to perform a systematic review and meta-analysis of antithrombotic regimens in post-TAVI patients with an indication for OAC. We compared OAC (VKA or DOAC) to OAC combined with single antiplatelet therapy (SAPT), either aspirin or clopidogrel, as antithrombotic treatment following TAVI. Endpoints included bleeding events, ischemic events, and all-cause mortality.

## METHODS

### Protocol

This study was designed according to the Preferred Reporting Items for Systematic Reviews and Meta-Analyses (PRISMA) statement (**Supplementary Table 1**).

**Table 1.**
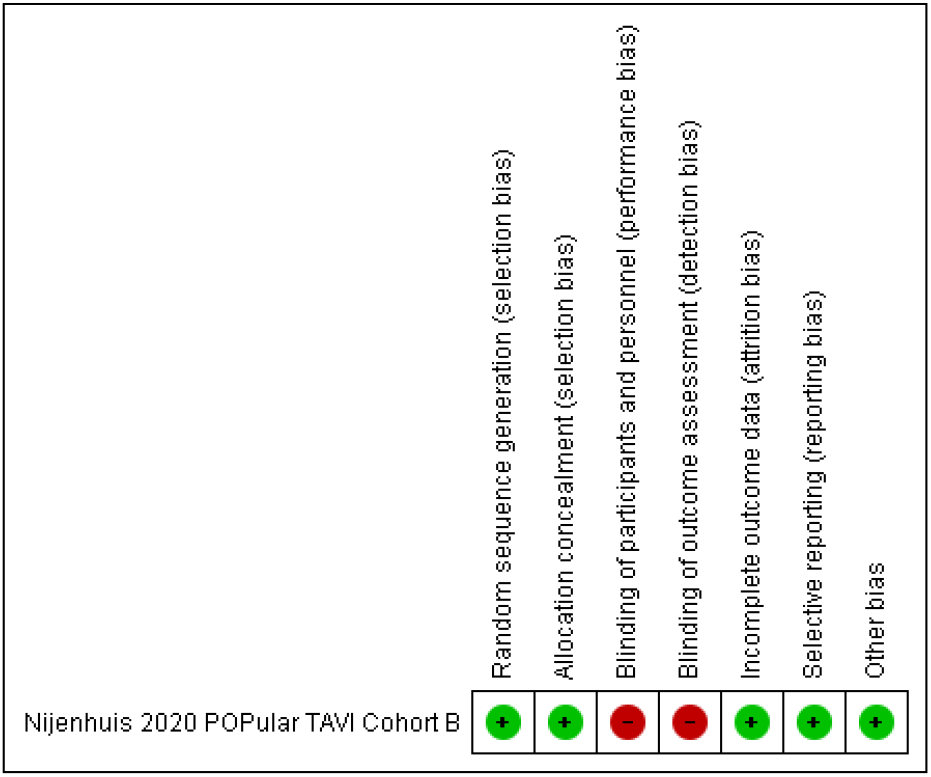
Risk of Bias Summary.

### Literature Search

We systematically searched PubMed, Embase, and the Cochrane Controlled Register of Trials (CENTRAL) in April 2020 for interventional and observational studies comparing OAC and OAC plus SAPT in patients undergoing TAVI who had an indication for anticoagulation. The search was limited by language (English, French, Portuguese, or Spanish) and type of subjects (human). No publication date limits were imposed. **Supplementary Figure 1** outlines the search strategy. Additional data were collected from randomized controlled trial (RCT) protocols. Patients or the public were not involved in the design, or conduct, or reporting, or dissemination plans of our research

**Figure 1.**
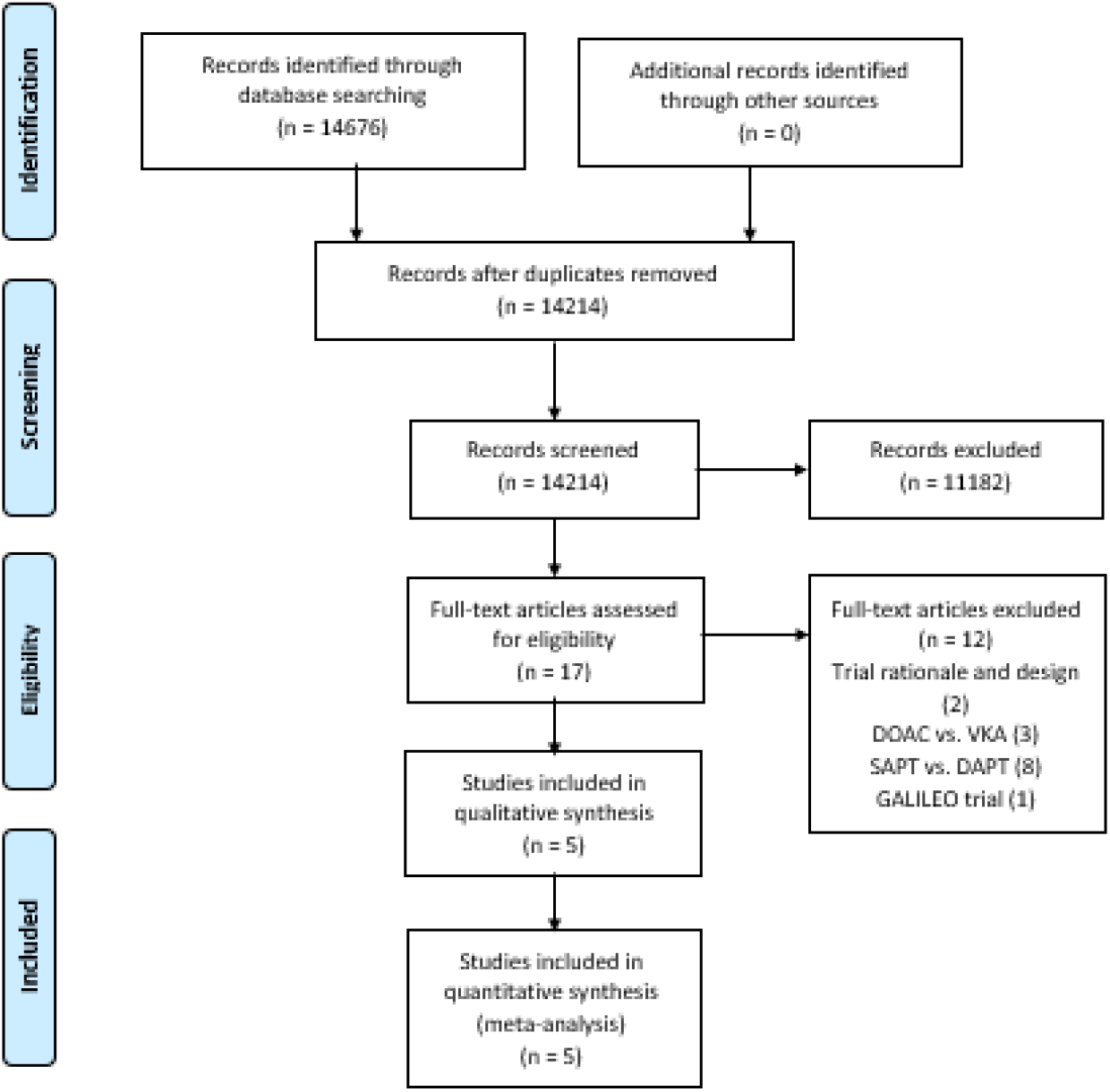
Flow diagram of literature search.

### Eligibility Criteria

The following criteria defined study eligibility: 1) interventional or observational studies comparing OAC with OAC plus SAPT in patients undergoing TAVI; 2) participants with symptomatic severe AS; 3) patients with an indication for long-term OAC; and 4) information on bleeding and ischemic complications during the follow-up period. We excluded series with fewer than 20 patients or without full-text article publications.

### Primary and Secondary Outcomes

The primary endpoints were severe bleeding, major bleeding, life-threatening bleeding, and intracranial bleeding. Severe bleeding included major and life-threatening events, which were defined according to the Valve Academic Research Consortium-2 (VARC-2) (12). Major bleeding is overt bleeding associated with a decrease in hemoglobin level of ≥ 3.0 g/dL or requiring transfusion of two or three units of whole blood/red blood cells (RBC), bleeding causing hospitalization or permanent injury, or bleeding requiring surgery and bleeding that does not meet criteria for life-threatening bleeding. Life-threatening bleeding is fatal bleeding or bleeding in a critical organ, such as intracranial, intraspinal, intraocular, or pericardial bleeding necessitating pericardiocentesis; intramuscular bleeding with compartment syndrome; bleeding causing hypovolemic shock or severe hypotension requiring vasopressors or surgery; or bleeding associated with a decrease in hemoglobin ≥ 5 g/dL or whole blood or packed RBC transfusion ≥ 4 units. VARC-2 definitions were used in all the included studies.

Secondary endpoints were all-cause mortality, myocardial infarction (MI), minor bleeding, and all-bleeding events. Stroke included ischemic and hemorrhagic stroke. All-bleeding included severe bleeding and minor bleeding events.

### Data Collection and Management

Two authors (GC, RT) systematically screened the titles and abstracts of publications retrieved to select studies that met the inclusion criteria. Next, they independently assessed the full texts of the studies for eligibility. Data was extracted concerning the study population, main demographics and baseline characteristics, interventions, and outcomes of interest. We analyzed studies with multiple sequenced publications and ensured no duplication of results and collection of the most recent data.

### Risk of Bias Assessment

The same two authors (GC, RT) independently assessed the risk of bias of the included articles, following the Cochrane Collaboration’s “Risk of bias” tool for RCTs and the Newcastle-Ottawa Scale for observational studies. RCTs were assessed as “low”, “high”, or “unclear” risk for the following biases: random sequence generation, allocation concealment, blinding of participants and personnel, blinding of outcome assessment, incomplete outcome data, selective reporting, and other bias. The quality assessment for each study is presented in the “Risk of Bias Summary” (**Table 1**) and the Newcastle-Ottawa Scale Summary (**Table 2**).

**Table 2.** Newcastle-Ottawa Scale Summary.

|  | Selection | Comparability | Outcome |
| --- | --- | --- | --- |
| D'Ascenzo 2017 | **** | * | ** |
| Geis 2017 | *** | * | ** |
| Abdul-Jawad<br>Altisent 2016 | **** | * | ** |
| Czerwińska-Jelonkiewicz 2016 | **** | — | ** |

### Statistical Analysis

We pooled dichotomous non-adjusted data using odds ratios (OR) to describe effect sizes using the Mantel-Haenszel procedure in a random effects model. Study heterogeneity was evaluated by funnel plots. The mean effect was considered significant if its 95% confidence interval (CI) did not include zero. Heterogeneity was assessed using the I^2^ statistic and assumed to be relevant if it exceeded 50%.

## RESULTS

### Search Results

Our search identified 14,726 articles, including one RCT (POPular-TAVI Cohort B). After removal of duplicates, we excluded 14,241 publications according to title and abstract evaluation, study type, and study population. Finally, five publications met all inclusion criteria, providing a total of 1318 patients for the meta-analysis (**Figure 1**). Study characteristics are shown in **Table 3**.

**Table 3.** Main Characteristics of Included Studies.

| Study | Regimen | Number of patients | Duration of follow-up, mnths | Post-TAVI Antithrombotic protocol | Type of valve |
| --- | --- | --- | --- | --- | --- |
| Nijenhuis et al. 2020 | OAC | 164 | 12 | OAC: VKA or DOAC | According to local protocol |
|  | OAC+SAPT | 162 |  | SAPT: Clopidogrel 3 months |  |
| D’Ascenzo et al.2017 | OAC | 105 | 6 | OAC: VKA | Edwards SAPIEN and SAPIEN XT prostheses |
|  | OAC+SAPT | 105 |  | SAPT: Aspirin 6 months |  |
| Geis et al. 2017 | OAC | 77 | 6 | OAC: VKA | Edwards SAPIEN 3, SAPIEN XT, Direct Flow Medical and CoreValve prostheses |
|  | OAC+SAPT | 41 |  | SAPT: Aspirin or Clopidogrel |  |
| Abdul-Jawad Altisent et al. 2016 | OAC | 101 | 13 | OAC: VKA | Edwards SAPIEN, SAPIEN 3, SAPIEN XT and CoreValve prosthesis |
|  | OAC+SAPT | 463 |  | SAPT: Aspirin or Clopidogrel |  |
| Czerwińska-Jelonkiewicz et al. 2016 | OAC | 38 | 12 | OAC: VKA | Edwards-Sapien/Sapien XT prosthesis |
|  | OAC+SAPT | 128 |  | SAPT: Aspirin or clopidogrel |  |
| OAC – Oral Anticoagulation; SAPT – Single Antiplatelet Therapy; VKA – Vitamin K Antagonis |  |  |  |  |  |

**Table 4.**
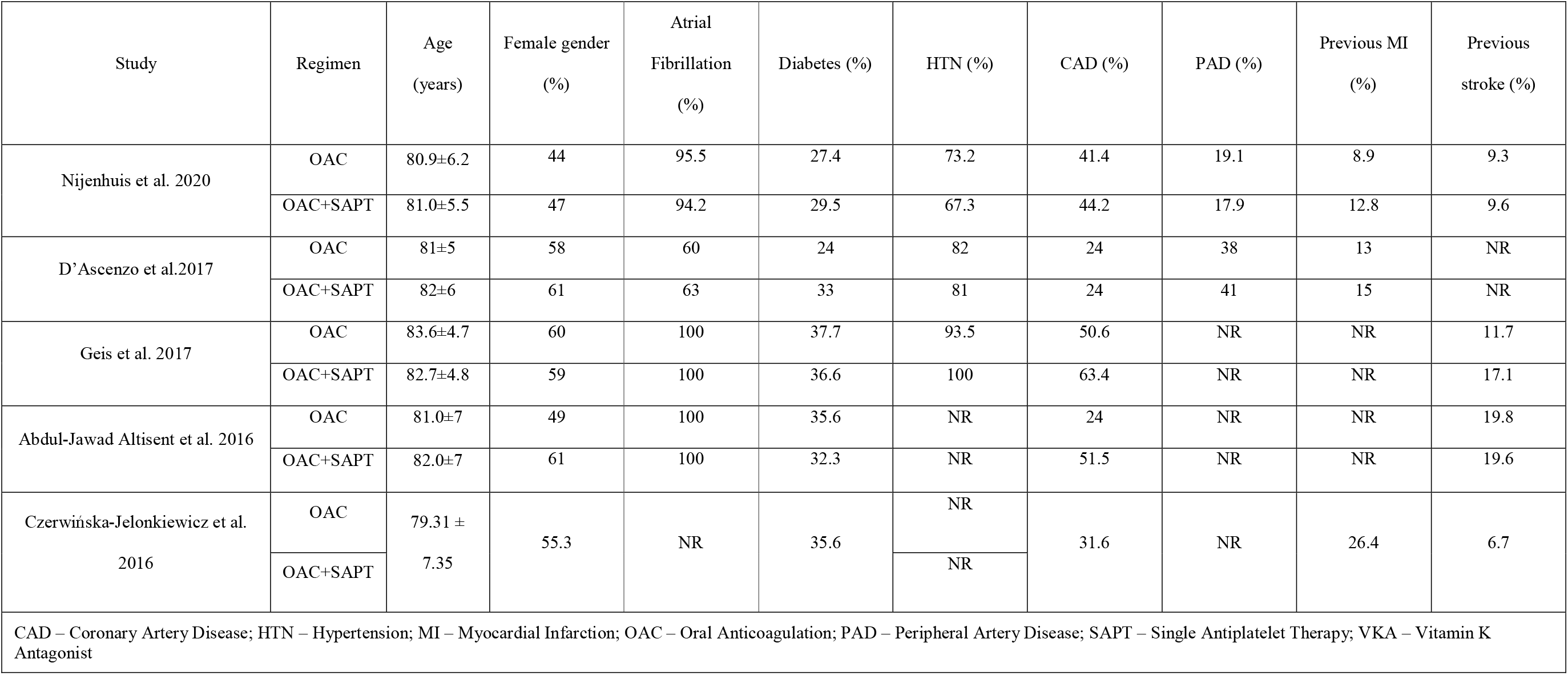
Clinical features in included studies.

### Primary Outcomes

Our meta-analysis revealed lower rates of severe bleeding (pooled OR 0.48 [0.34, 0.69], P<0.01, I^2^=0%) (**Figure 2**) and major bleeding (pooled OR 0.55 [0.36, 0.86], P<0.01, I^2^=0%) in the OAC group than in the OAC-plus-SAPT group. There was a nonsignificant trend towards reduced life-threatening bleeding events favoring the OAC group (pooled OR 0.54 [0.27, 1.08], P=0.08, I^2^=11%), but similar rates of intracranial bleeding were observed in the groups (pooled OR 1.57 [0.39, 6.24], P=0.52, I^2^=12%). No significant heterogeneity or selection/publication bias was identified (**Supplementary Figures 2**).

**Figure 2.**
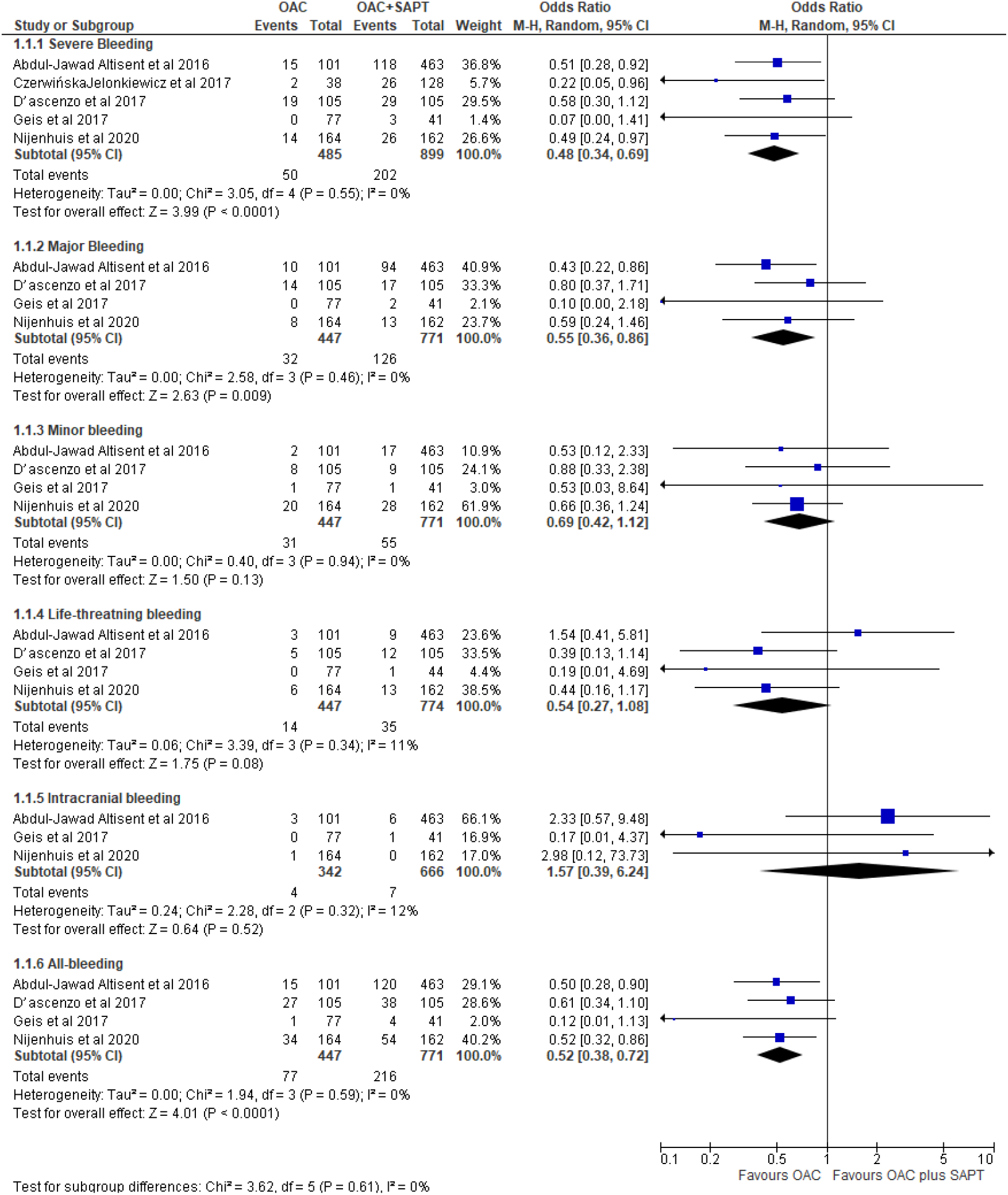
Forest Plot of Bleeding Events Comparing OAC versus OAC plus SAPT.

### Secondary Endpoints

There was no difference between the OAC and OAC-plus-SAPT groups in terms of stroke prevention after TAVI (pooled OR 0.98 [0.57, 1.70], P=0.58, I^2^=0%) (**Figure 3**). There were also similar rates of all-cause mortality (pooled OR 1.04 [0.75, 1.42], P=0.83, I^2^=0%) (**Figure 4**) and MI (pooled OR 0.36 [0.04, 3.07], P=0.35, I^2^=16%).

**Figure 3.**
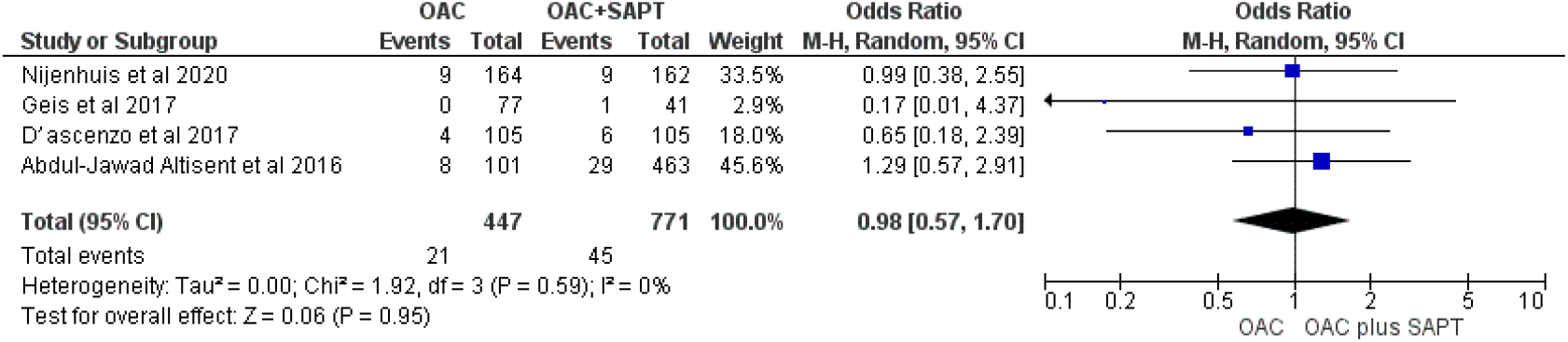
Forest Plot of Stroke Comparing OAC versus OAC plus SAPT.

**Figure 4.**
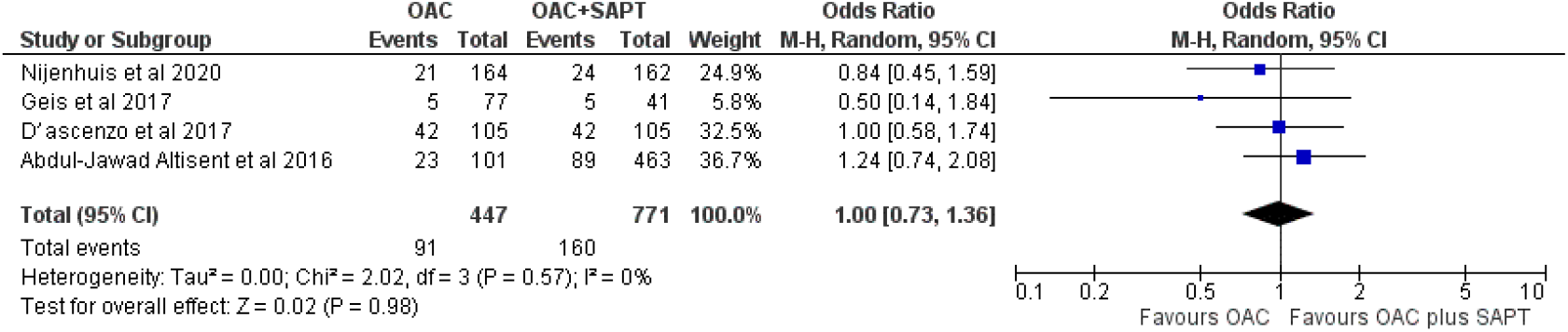
Forest Plot of All-Cause Mortality Comparing OAC versus OAC plus SAPT.

Additionally, the composite endpoint of all-bleeding events was lower in the OAC group than in the OAC-plus-SAPT group (pooled OR 0.52 [0.38, 0.72], P<0.01, I^2^=0%) (**Figure 1**). Minor bleeding events were similar between groups (pooled OR 0.68 [0.42, 1.11], P=0.13, I^2^=0%). No selection or publication bias was identified by funnel plots (**Supplementary Figures 2-4**).

## DISCUSSION

According to our systematic review and meta-analysis, for patients requiring long-term OAC after TAVI, the use of double anti-thrombotic therapy, compared to the use of OAC alone, significantly increased the risk of bleeding, including major events. No significant differences were noted regarding ischemic events or all-cause mortality.

Antiplatelet therapy is combined with anticoagulation to prevent thromboembolic complications before valve endothelialization is completed, but this is a controversial practice in interventional cardiology. American and European guidelines on post-TAVI antithrombotic regimens consider the flow conditions of a bioprosthetic valve similar to a coronary stent(4,5,7,8,13). Many patients with AF have concomitant coronary artery disease (CAD) and undergo percutaneous coronary intervention (PCI) with stenting. These patients often receive aspirin therapy in addition to OAC. This combination reduces ischemic and thrombotic events but increases the risk of major bleeding by 2-to 3-fold (14,15).

The risk of bleeding with combined OAC plus antiplatelet therapy is related to the degree of anticoagulation, patient characteristics, and comorbidities. The patients in our analysis had a high-risk profile, so our pooled results do not apply to younger or intermediate-to low-risk patients(16,17).

### Post-TAVI Stroke Rate

A recent analysis of the Transcatheter Valve Therapy Registry reported a stable 30-day stroke risk after TAVI in the first five years after the procedure’s approval by the United States Food and Drug Administration(18). Improvements in TAVI technology and operator experience positively impacted major post-TAVI complications such as bleeding, vascular injury, and mortality but did not translate into a reduced stroke rate(19).

A potential strategy to prevent embolic events during TAVI is the use of embolic protection devices. A recent meta-analysis suggested that the use of such devices appears to be associated with a nonsignificant trend towards reduced death or stroke(20). Future evidence related to this issue could lead to a significant decrease in stroke risk after TAVI.

A left atrial appendage thrombus is a potential source of post-TAVI stroke for AF patients(21). The ongoing WATCH-TAVR trial is a prospective, multicenter, RCT (NCT03173534) that is evaluating the safety and effectiveness of left atrial appendage occlusion for the prevention of stroke and bleeding hazards in patients with AF undergoing TAVI.

In most TAVI studies, baseline populations include patients with AS who were rendered unsuitable for surgical aortic replacement (SAVR), meaning they are elderly patients with high operative risk(22). This subset of patients commonly has several comorbidities that impact embolic and bleeding events(23). Two recent trials assessing the value of TAVI in low-risk patients demonstrated non-inferiority of the technique when compared with SAVR for major outcomes, which will likely expand TAVI indications(24,25). One of these trials reported no mortality or stroke events at 30 days, demonstrating the safety of the procedure(24). Nevertheless, leaflet thrombosis, a potential risk factor for stroke and transient ischemic attacks, was reported in a minority of patients, although there was no association with clinical events. Therefore, long-term follow-up of these patients will be important for understanding the impact of subclinical leaflet thrombosis and determining the best antithrombotic management for this subset of patients(26).

An important confounding variable in our study is PCI prior to TAVI. Concomitant antiplatelet therapy is recommended in patients with unstable CAD or following stenting, with triple antithrombotic therapy as a reasonable approach in this specific setting. The POPular-TAVI trial excluded patients with drug-eluting stent implantation within 3 months and bare-metal stent implantation within 1 month prior to TAVI(11). It did not show a significant difference in the prevalence of coronary disease or previous MI between the two treatment arms. However, the same findings were not true in the rest of the included studies.

### Future: VKA versus DOACs after TAVI?

DOACs were developed to overcome the limitations of VKA: they have demonstrated a consistently favorable efficacy/safety profile for the prevention of thromboembolic complications of AF and are widely used in clinical practice(27,28). In a meta-analysis of four AF trials comparing DOACs with VKA, the rates of intracranial hemorrhage were nearly halved with DOACs and ischemic stroke and mortality were also significantly reduced(29). These conclusions were similar for patients with non-valvular AF and concomitant aspirin therapy(30).

The broad use of DOACs in clinical practice is reflected in the original POPular-TAVI trial, in which DOAC use was an exclusion criteria, but it was not fully eliminated from the ultimate patient sample(31). Furthermore, a prespecified subgroup analysis of this trial showed a possible benefit of DOACs over VKA regarding bleeding or ischemic outcomes(11). Nevertheless, most of the available data on DOACs after TAVI are registry-based; they show an improved safety profile with no difference in ischemic outcomes when compared with VKA(32). Additionally, in the RESOLVE and SAVORY registries, there were no significant differences in reduced leaflet motion between DOAC and warfarin; both appeared better than no anticoagulation(33). A substudy of the ENGAGE AF-TIMI 48 trial, comprising 91 patients with previous transcatheter or surgical bioprosthetic valve implantation and AF demonstrated a significant reduction of major bleeding with low-dose (30 mg) edoxaban compared with warfarin(33). Thus, DOAC-based antithrombotic regimens seem a valid treatment option in patients undergoing TAVI who require long-term OAC. The ongoing ENVISAGE-TAVI AF (NCT02943785) is expected to reveal more insight into this issue.

## Limitations

The studies included in our meta-analysis primarily compared VKA-based treatments, which is a major limitation of this study. Another important limitation of our meta-analysis is the paucity of RCTs on this issue. In fact, most studies included were observational, which increases the risk of bias and limits the strength of our results.

Minor bleeding was defined differently in the included studies and between centers in the POPular-TAVI trial. Substantial heterogeneity exists between studies in terms of design and type and duration of antiplatelet/antithrombotic therapy used. Nevertheless, the heterogeneity did not affect our analysis, and all I^2^ values were < 50%. Additionally, most of the studies did not report the incidence of MI and some did not differentiate type of stroke.

Another limitation of our analysis is that no study distinguished forms of intracranial bleeding other than hemorrhagic stroke, which could underrepresent the real incidence of this outcome. Furthermore, antiplatelet therapy duration and specific antiplatelet drug used in the antithrombotic regimen were not reported or differed among studies. Consequently, these comparisons should be interpreted with caution.

## CONCLUSION

Our pooled analysis suggests that for patients with an indication for long-term OAC after TAVI, the use of double anti-thrombotic therapy (OAC with SAPT), compared to the use of OAC alone, increased the risk of bleeding without reducing all-cause mortality and cerebrovascular events.

## Data Availability

All data referred in the manuscript is available to the general public

## Abbreviation index

AF: Atrial Fibrillation
AS: Aortic Stenosis
CAD: Coronary Artery Disease
HTN: Hypertension
MI: Myocardial Infarction
NR: Non-Reported
OAC: Oral Anticoagulation
PAD: Peripheral Artery Disease
PRISMA: Preferred Reporting Items for Systematic Reviews and Meta-Analyses
PCI: Percutaneous Coronary Intervention
RCT: Randomized Controlled Trial
SAPT: Single Antiplatelet Therapy
TAVI: Transcatheter Aortic-Valve Implantation
VARC-2: Valve Academic Research Consortium-2
VKA: Vitamin K Antagonist

